# A Virtual Reality-Based Assessment Tool for Apathy

**DOI:** 10.1101/2025.05.26.25328377

**Authors:** Daniel Harlev, Ramit Ravona-Springer, Or Koren, Shlomit Zorani, Meir Plotnik, Noham Wolpe

## Abstract

**Importance:** Apathy is a common and disabling syndrome in aging and neurodegenerative disease. Existing assessment tools rely on subjective reporting, limiting their accuracy in cognitively impaired individuals or in individuals with otherwise impaired insight.

**Objective:** To identify behavioral subgroups relevant to apathy using gaze-based metrics in an emotionally salient virtual reality (VR) setting, and to characterize their clinical and physiological profiles.

**Design, Setting, and Participants:** Cross-sectional study of 85 older adults (mean age 74 ± 5.7 years) with varying cognitive abilities, recruited from a memory clinic. Participants completed a naturalistic VR task while gaze and physiological signals were recorded.

**Exposures:** Positive, aversive, and neutral images were presented within a 3D VR environment. Gaze features included time to first fixation, total fixation duration, and fixation variability. Physiological signals included heart rate variability (HRV), galvanic skin response, and respiration.

**Main Outcomes and Measures:** Unsupervised clustering (K-means) was applied to gaze metrics to derive behavioral engagement profiles. Between-group differences in apathy severity (LARS), cognitive function (MoCA), and physiological reactivity were examined using linear mixed-effects models.

**Results:** Two distinct behavioral subgroups emerged: “engagers” (n = 48) and “non-engagers” (n = 37). Non-engagers were older, showed higher apathy scores, and had lower MoCA scores. Physiologically, they exhibited lower baseline HRV but showed a selective increase in HRV during exposure to positive stimuli. No group differences were observed in response to aversive stimuli or in other physiological signals.

**Conclusions and Relevance:** This study introduces a novel, nonverbal approach for identifying apathy through naturalistic gaze behavior in VR. The findings highlight a physiologically responsive yet behaviorally disengaged subgroup, suggesting that emotional reactivity may persist even when outward engagement is diminished. Gaze-based phenotyping in immersive environments may offer a scalable tool for detecting motivational impairments in aging and can be further tested in individuals with poor insight to inform future personalized assessment strategies.

## Introduction

Apathy is a prevalent and debilitating neuropsychiatric syndrome in late life, often manifesting as diminished motivation, reduced goal-directed behavior, and emotional indifference (Miller et al., 2021). It has been observed across a wide range of neurological and psychiatric conditions, including Alzheimer’s disease, Parkinson’s disease, and late-life depression (Harlev, Singer, et al., 2025; Husain & Roiser, 2018). Importantly, apathy itself, irrespective of one’s affective or cognitive states, is associated with poor functional outcomes, increased caregiver burden, and accelerated disease progression (Lanctôt et al., 2023; Steffens et al., 2022). Apathy has also been associated with increased risk for cognitive decline and dementia, suggesting its potential role as an early behavioral marker (Bock et al., 2020; Marshall et al., 2013).

The tools most commonly used to assess apathy are questionnaires administered to patients or their caregivers, such as the Lille Apathy Rating Scale (LARS). However, these questionnaires depend on subjective reports (Mohammad et al., 2018). While these instruments are practical in clinical settings, they are prone to reporting biases, and poor accuracy in individuals with reduced insight such as cognitively impaired populations (Radakovic et al., 2015).

Emerging models of psychopathology, such as RDoC, emphasize the need for biologically informed assessment tools that capture dimensions of behavior not bounded by conventional diagnostic categories (Thant & Yager, 2019). This conceptual shift motivates the search for novel and objective approaches that can reveal biologically meaningful behavioral phenotypes relevant to apathy.

One such method is virtual reality (VR), which enables the study of behavior in immersive, ecologically valid environments (Parsons, 2015). By embedding emotionally salient stimuli within dynamic and realistic settings, VR enables to observe spontaneous, task-free behavior that more closely mirrors ‘naturalistic’ or real-world functioning (Chirico & Gaggioli, 2019). This approach is particularly useful when combined with continuous nonverbal behavioural measurement techniques, such as eye gaze, and allows for the concurrent recording of physiological measurements. Passive gaze-based metrics, including time to first fixation (TTFF), total fixation duration (TFD), and gaze variability, can serve as proxies for attention, emotional reactivity, and motivational engagement (Kim, 2024). Concurrently, physiological indices, such as heart rate variability (HRV), galvanic skin response (GSR), and respiration can provide insights into autonomic regulation in response to emotional stimuli (Appelhans & Luecken, 2006; Civitello et al., 2014; Egger et al., 2019). Together, these measures offer an opportunity to study apathy-related processes through objective, temporally precise signals that are not dependent on subjective verbal reporting—an approach which is particularly valuable in individuals with cognitive impairment and neuropsychiatric disorders.

Here, we developed a novel VR-based emotional viewing task designed to objectively measure apathy in clinical settings which we recently validated (RavonaLSpringer et al., 2025). Our principal objective was to explore whether this VR tool can provide an objective measure of behavioral engagement that is consistent with clinical scales but offers additional clinical value by not having to rely on self-report. To this end, we applied an unsupervised clustering approach to derive behavioral groupings from gaze data collected during naturalistic viewing behaviour. Our goal was to test whether these data-driven clusters would align with clinically relevant features, including apathy and cognition, as well as with physiological reactivity to emotional stimuli. This approach would enable the discovery of implicit behavioral phenotypes that may not be captured by traditional symptom-based classifications.

## Methods

### Participants and clinical assessment

Participants were recruited from the Geriatric Psychiatry and Memory Unit at Sheba Medical Center, Israel (IRB approval reference 4436-17). Inclusion criteria included participants aged 60–90 years, with mild cognitive impairment to mild dementia, based on clinical assessment and neurocognitive testing. Exclusion criteria included severe dementia (Mini-Mental State Examination score ≤ 12), physical health conditions affecting compliance, or significant mental health disorders other than apathy or depression. Of the 97 participants initially recruited, 85 were included in the analysis, with 12 participants excluded due to incomplete or unusable eye gaze data resulting from calibration failures or technical issues during the VR task. Self-reported apathy was assessed using the short version of the LARS, which is a structured interview administered to the patient, yielding scores from −36 to +36, with higher scores reflecting greater apathy severity (Sockeel et al., 2006). Self-reported depressive symptoms were assessed using the 15-item Geriatric Depression Scale (GDS-15), with scores ranging from 0 to 15, where higher scores correspond to greater depression severity (Bijl et al., 2006). Cognitive function was evaluated using the Montreal Cognitive Assessment (MoCA), which is a screening tool with a score range of 0 to 30, where higher scores indicate better cognitive performance (Nasreddine et al., 2005). All demographic and clinical information is summarized in Table 1.

**Table 1.**
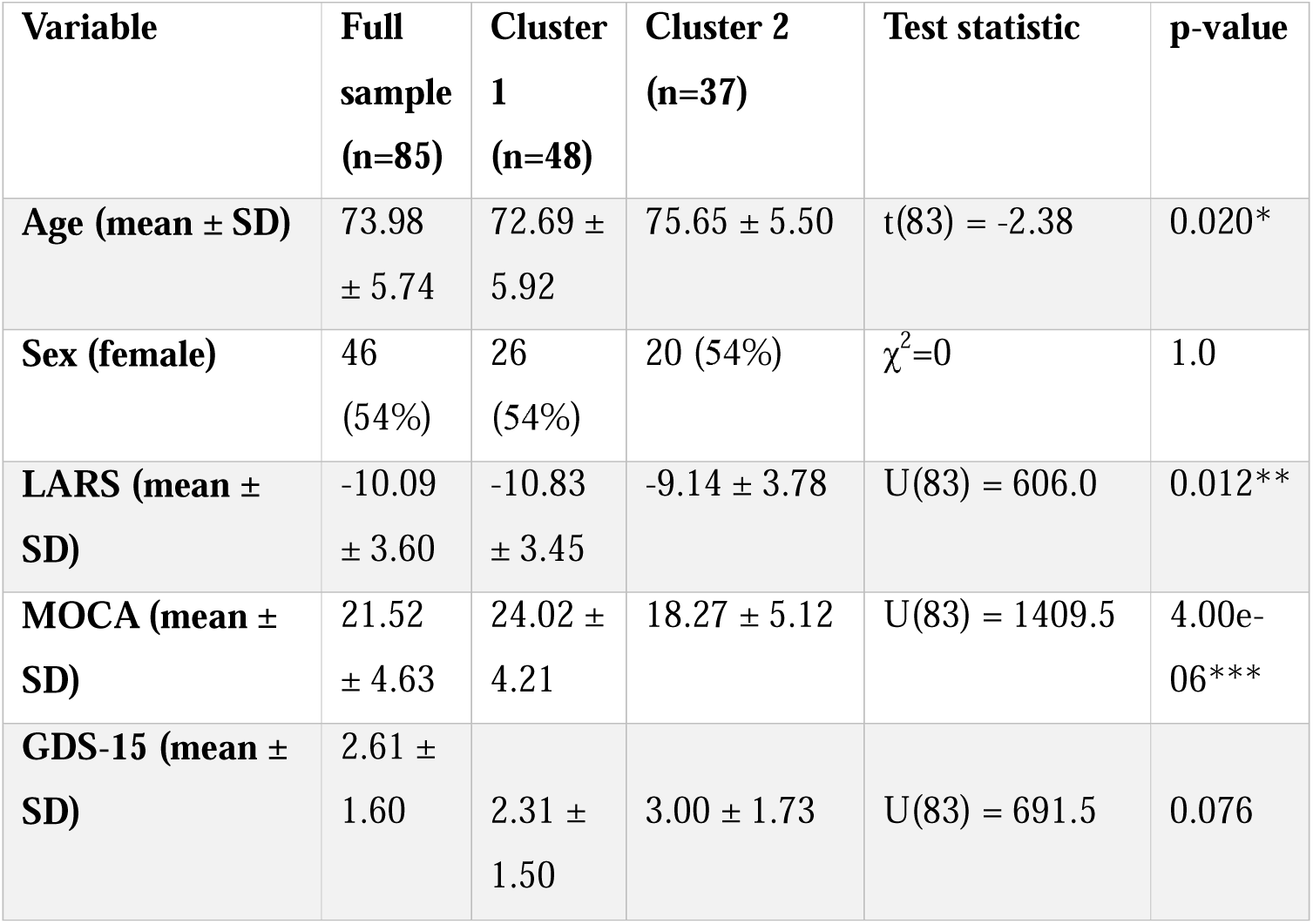
Demographics and clinical measures for the full sample and each cluster. Demographic, cognitive, and affective characteristics of participants are presented for the full sample (N=85), Cluster 1 (“engagers; n=48), and Cluster 2 (“non-engagers; n=37). Data include means (±SD) for continuous variables and proportions for categorical variables. Significant differences between clusters were determined using independent t-tests (for normally distributed continuous variables), Mann-Whitney U tests (for non-normally distributed continuous variables), and chi-square tests (for categorical variables).

### Virtual reality task and stimuli

Participants viewed emotionally salient stimuli within a virtual urban park environment using a head-mounted VR display. Positive, negative, and neutral images were embedded on billboards and moving vehicles, presented over 36 repetitions or trials. Each stimulus was displayed for 10 seconds, with free-viewing instructions and a fixed interstimulus interval of 30 seconds. Emotionally positive stimuli were personalized images of grandchildren, while negative and neutral stimuli were drawn from validated image databases (full protocol and example screenshots provided in the Supplementary Methods).

### Gaze and physiological measures

Eye-tracking data were collected continuously throughout the VR session and used to compute three key gaze metrics: time to first fixation (TTFF), total fixation duration (TFD), and intra-individual fixation variability. Concurrent physiological recordings included heart rate (HR), respiration (rate and depth), galvanic skin response (GSR), and heart rate variability (HRV). Baseline and post-stimulus reactivity values were defined over 10-second windows before and after stimulus onset. Preprocessing procedures and signal derivation steps are described in full in the Supplementary Methods.

### Unsupervised Clustering Analysis

K-means clustering was applied to gaze features (TTFF, TFD, and variability) to identify behavioral engagement profiles. The optimal number of clusters was selected based on silhouette and Davies-Bouldin metrics. Cluster robustness was tested through noise-perturbation and ceiling-trial exclusion analyses, with assignment consistency quantified using adjusted mutual information and adjusted Rand index (full procedures detailed in Supplementary Methods).

Following gaze-based clustering, we tested for individual differences between participants in the identified clusters in terms of age, sex, cognitive function (MoCA), apathy (LARS) and depression (GDS-15), using Mann-Whitney U tests. To explore whether the clustering captured meaningful differences in autonomic reactivity to different emotional stimuli, we conducted a set of Linear Mixed Models (LMMs). First, we examined physiological measures across clusters, with cluster assignment and age as fixed effects, and individual intercepts for each participant. Separate models were constructed for the six physiological measures, with *post hoc* analyses stratified by stimulus type (negative, neutral, positive) to investigate differential physiological responses.

All analyses were conducted in Python (v3.10) using the statsmodels and scikit-learn packages (Pedregosa et al., 2011). Significance was set at α = 0.05, with False Discovery Rate (FDR) correction applied for multiple comparisons using the Benjamini-Hochberg procedure (Benjamini & Yekutieli, 2001).

## Results

Participant demographic and clinical characteristics are presented in Table 1.

### Behavioral clustering

We performed K-means clustering on three gaze metrics: TTFF, TFD and FVI. To determine the optimal number of clusters, we examined two widely used internal validation metrics: the Silhouette Score and the Davies-Bouldin Index (Arbelaitz et al., 2013). Both indices consistently indicated that two clusters (k=2) provided the best separation between participants (Fig. 1A-B). Compared to Cluster 2 (“non-engagers”; n=37), Cluster 1 (“engagers”; n=48) showed shorter TTFF, longer TFD, and lower FVI values (Fig. 2).

**Figure 1.**
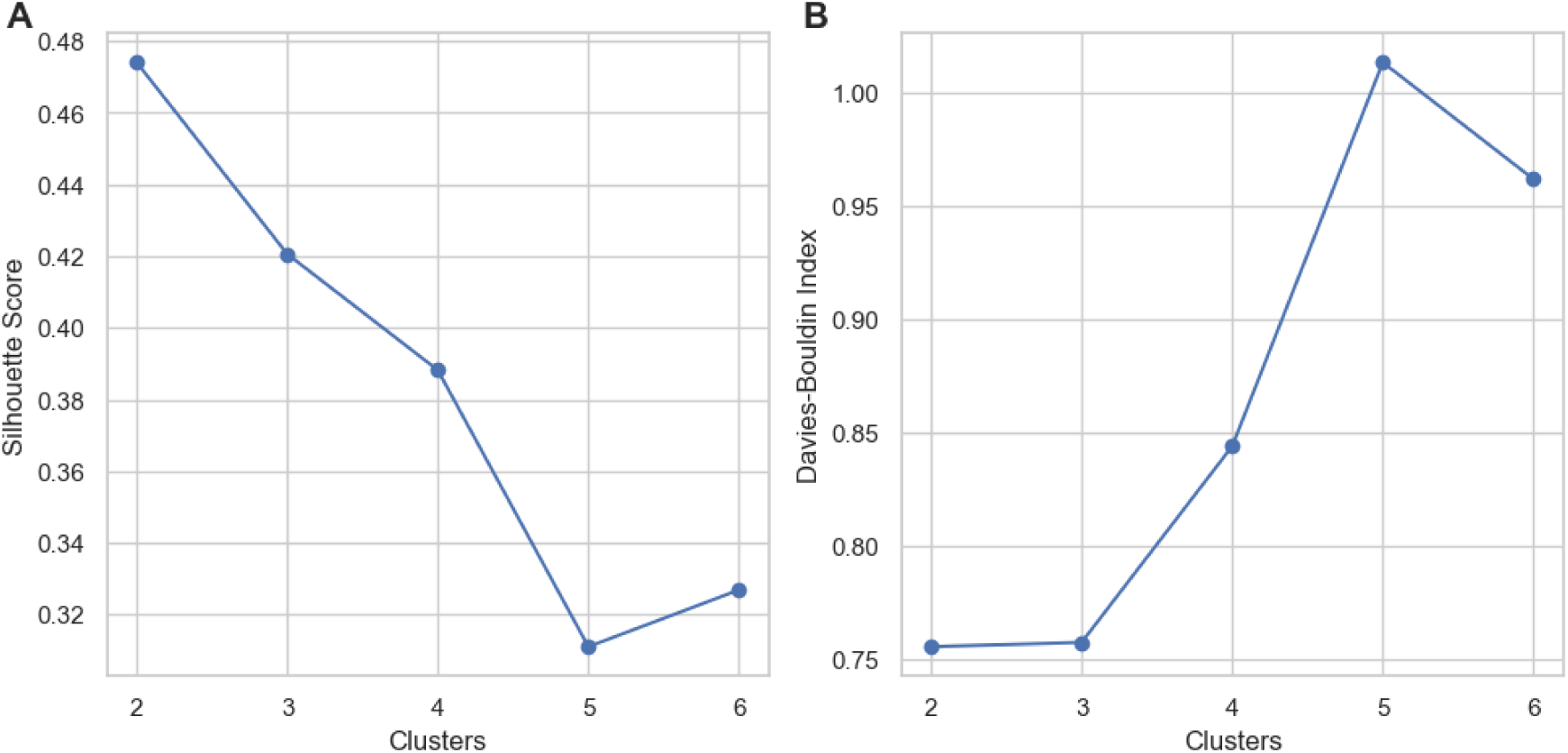
Cluster validation metrics for unsupervised gaze-based classification. (A) Silhouette Score plotted across a range of cluster solutions (k = 2 to 6). The Silhouette Score reflects both the cohesion within clusters and the separation between them, with higher values indicating better-defined and more distinct clusters. (B) Same as (A), but for Davies-Bouldin Index, which quantifies the average similarity between each cluster and its most similar peer. Lower values indicate more compact and well-separated cluster configurations.

**Figure 2:**
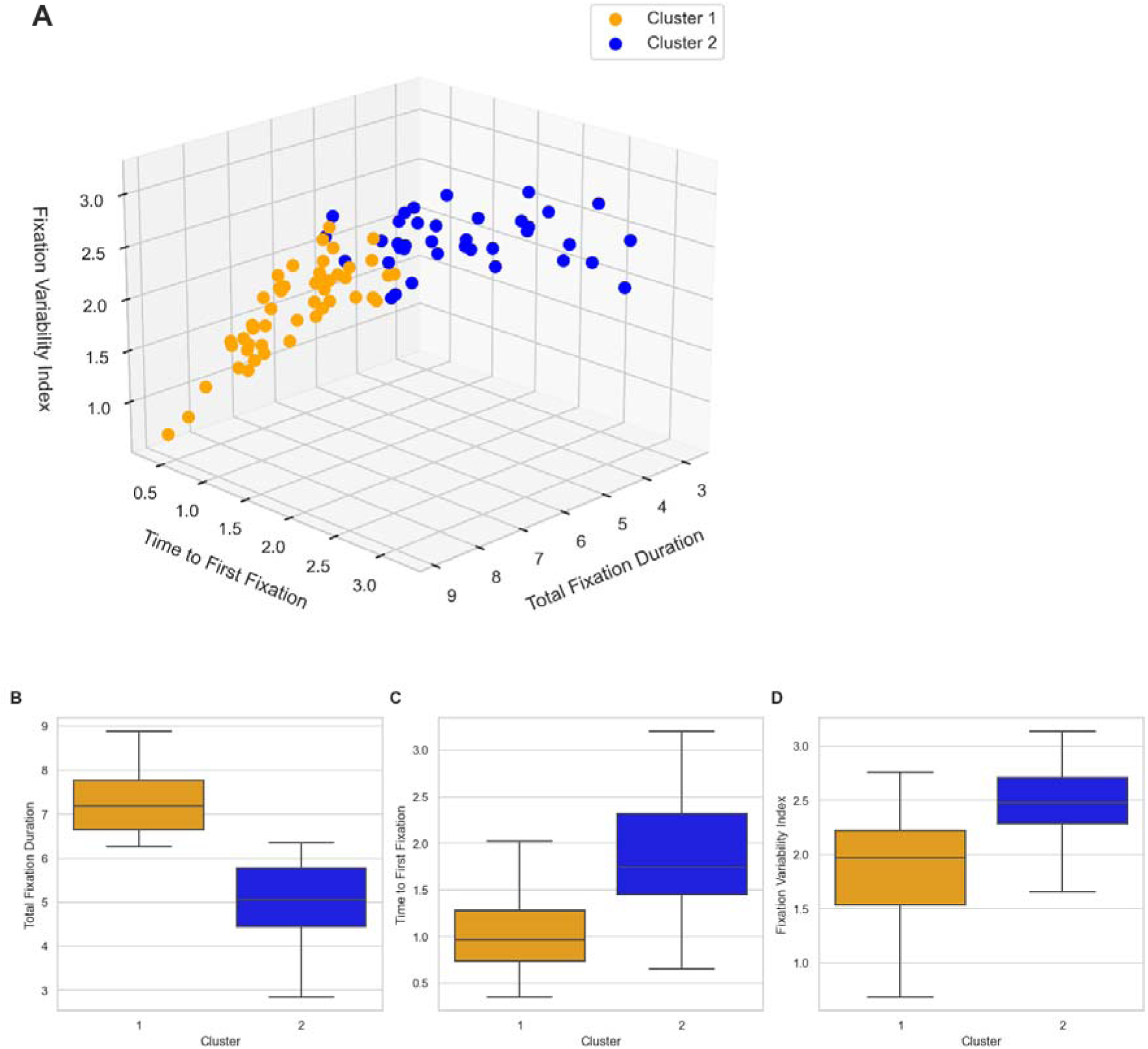
K-Means clustering of gaze-based metrics and group-level comparisons. (A) Three-dimensional scatterplot illustrating participant distribution based on Total Fixation Duration (X-axis), Time to First Fixation (Y-axis), and the Fixation Variability Index (Z-axis). Markers are colored by group assignment based on K-means clustering. (B–D) Box plots comparing gaze metrics between clusters for Total Fixation Duration (B), Time to First Fixation(C), and Fixation Variability Index (D).

To assess clustering robustness, we computed the AMI across 1000 iterations with Gaussian perturbation. The resulting AMI (*M* = 0.777, *SD* = 0.086) suggests that the clustering captured a robust structure with high consistency across resampled datasets. Moreover, excluding trials where [TTFF + TFD] ≥ 9.9 seconds (605/18.24% trials) yielded identical participant assignments, as indicated by an ARI of 1.00. These results suggest a robust cluster structure which was not unaffected by edge-case trials.

### Cluster clinical profiles

The clusters differed across several clinical domains, as summarized in Table 1 (All *p*-values were FDR-corrected for multiple comparisons). As shown in Table 1, non-engagers were significantly older than engagers (*t*(83) = −2.38, *p* = 0.020). Differences in apathy levels were also observed, with non-engagers showing higher LARS scores (*U*(83) = 606.0, *p* = 0.018). Additionally, MoCA scores were significantly lower for non-engagers (*U*(83) = 1409.5, *p* <0.001). No significant difference was found in GDS-15 total scores, but a trend toward higher GDS-15 scores in Cluster 2 was observed (*U*(83) = 691.5, *p* = 0.077).

To determine whether self-report apathy independently predicted behavioral group membership, we conducted logistic regression analyses with LARS scores as the main predictor. LARS score significantly predicted group assignment when entered alone (β = −0.119, *p* = 0.041), and remained significant after adjusting for age (β = −0.117, *p* = 0.045). However, when controlling for cognitive performance (MoCA), the effect of apathy was no longer significant (β = −0.069, *p* = 0.276), while MoCA remained a strong predictor (β = 0.191, *p* < 0.001). Variance inflation factors were all <1.2, indicating reasonable multicollinearity. These results suggest that apathy and cognition contribute overlapping, yet also distinct variance to behavioral disengagement seen in the VR.

### Cluster physiological profiles

To further investigate the relevance of these clusters, we examined whether physiological responses to VR stimuli differed between the groups. Linear mixed models (LMMs) revealed a significant cluster difference in HRV reactivity during VR exposure, with higher values observed in the non-engager group (β *=* 0.205, *SE* = 3.182, *p* = 0.012). No significant differences between clusters were found for GSR, heart rate, respiration rate, or respiration depth (Table 2).

**Table 2.** Results of linear mixed-effects models examining the effect of cluster assignment on physiological response measures. . Each model included cluster as the fixed effect of interest, with a random intercept for each participant and age as a covariate. Coefficients are standardized based on the residual standard deviation of the outcome variable to allow comparability across features. Galvanic skin response (GSR) is measured in microsiemens; heart rate is measured in beats per minute; Heart rate variability (HRV) is measured as the root mean square of successive differences; Respiration rate and depth represent breathing frequency and amplitude, respectively. Significant results are indicated by *p < 0.05, **p < 0.01, ***p < 0.001.

| Feature | Cluster estimate<br>(standardized) | SE | p-value |
| --- | --- | --- | --- |
| GSR | -0.155 | 10.817 | 0.233 |
| Heart rate | -0.181 | 0.311 | 0.160 |
| HRV | 0.205 | 3.182 | 0.012** |
| Respiration rate | -0.016 | 1.464 | 0.851 |
| Respiration depth | 0.059 | 2.688 | 0.559 |

We next explored whether cluster differences in HRV reactivity were specifically driven by certain stimulus valence. A significant interaction emerged between cluster and stimulus valence for predicting HRV. This interaction was specifically driven by the positive condition, where participants in the non-engager cluster showed significantly higher HRV reactivity (β = 0.240, *SE* = 3.726, *p* = 0.016; Fig. 3). These results indicate that emotionally meaningful stimuli can elicit autonomic responses in the non-engager group, despite their reduced gaze engagement. As the analysis focused on changes in HRV relative to baseline, we explored whether these results were driven by differences in pre-stimulus baseline HRV values. This revealed that participants in the non-engager cluster exhibited significantly lower baseline HRV compared to the engager cluster (*t*(83) = −2.26, *p* = 0.024). Together, increased reactivity in the non-engager cluster can be partly explained by their lower baseline HRV, which could allow greater physiological modulation in response to positive stimuli.

**Figure 3.**
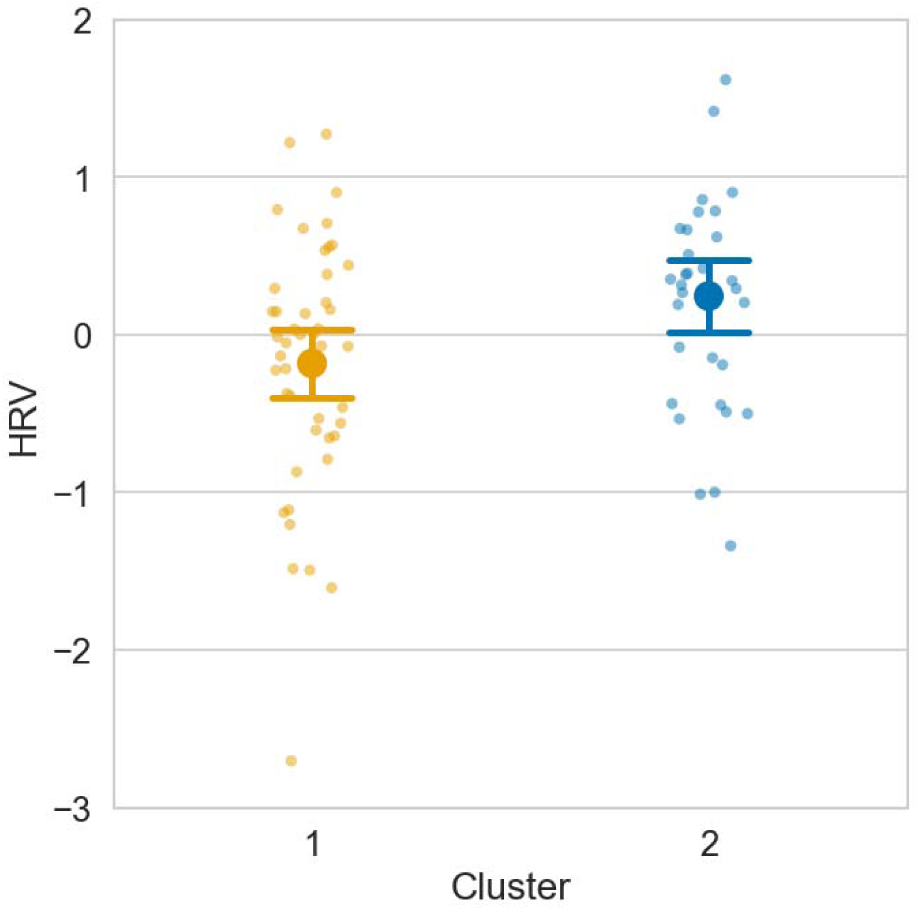
Age-adjusted heart rate variability (HRV) differences by behavioral cluster in the positive stimuli. Mean differences between HRV in response to positive stimuli relative to baseline, with 95% confidence intervals (CI). Values represent age-adjusted residuals obtained from linear regression models controlling for chronological age. Markers are colored by group assignment based on K-means clustering. Error bars indicate ±2 standard error of the mean.

## Discussion

This study identified two distinct behavioral engagement subtypes in older adults with varying levels of apathy and cognitive function, based on gaze pattern in a VR setup designed to assess apathy (RavonaLSpringer et al., 2025). Using an unsupervised clustering approach, we uncovered groups that were not predefined by symptom scores, yet differed systematically in self-report apathy scores, cognitive function and age. These clusters also showed divergent physiological profiles in HRV.

Our approach complements conventional measures of apathy that rely on subjective ratings. Apathy in aging is increasingly recognized as a multidimensional construct with strong neurocognitive underpinnings—particularly in executive function and attentional control— rather than a mere emotional flattening or depressive symptom (Lanctôt et al., 2023). This reframing has critical implications, as apathy may represent one of the earliest behavioral indicators of neurodegenerative processes (Bock et al., 2020). Our findings echoed this strong link: although apathy severity (LARS) was associated with gaze-based cluster assignment, this effect did not remain significant after adjusting for cognitive performance (MoCA), highlighting the partial overlap between affective disengagement and cognitive decline in late life. This result supports the view that apathy reflects impaired goal-directed cognition rather than reduced emotional experience, consistent with its role as a marker of cognitive vulnerability (Harlev, Singer, et al., 2025; Harlev, Vituri, et al., 2025; Lanctôt et al., 2023). Importantly, these insights emerged neither from clinical judgment nor self-reports, but from objective, spontaneous behavior in a naturalistic VR setting. This reinforces the potential of passive digital phenotyping in psychiatry in general, and in geriatric psychiatry in particular (Alfalahi et al., 2023; Bufano et al., 2023).

The clearest physiological difference between these behavioral subtypes was found in HRV, reflecting autonomic reactivity and readiness to engage with salient stimuli (Beauchaine & Thayer, 2015). The interpretation of this pattern goes beyond simple group-level differences. Rather than suggesting that one group was physiologically “blunted” while the other was “intact,” the results pointed toward a more nuanced, context-dependent form of disengagement. Compared to the behaviorally ‘engager’ group, individuals in the ‘non-engager’ group appeared physiologically reactivity in general. However, this pattern did not hold across all conditions. Specifically, under the positive condition, the non-engager group showed a marked increase in HRV, despite their otherwise reduced behavioral reactivity.

Notably, this response reflects a preserved physiological sensitivity to socially meaningful cues. One possible explanation is that non-engagers, who exhibited lower baseline HRV, may have retained a greater physiological capacity for modulation in response to meaningful social stimuli. This greater “physiological space” for reactivity might enable stronger autonomic responses under affiliative conditions. In contrast, participants with higher baseline HRV may have shown limited modulation due to vagal saturation (Beauchaine & Thayer, 2015).

The selective increase in HRV during the positive condition may reflect an age-related positivity bias—a tendency of older adults to prioritize emotionally meaningful stimuli, particularly those with social or affiliative value (Kennedy & Mather, 2024; Mather & Carstensen, 2003). While often considered an adaptive motivational shift, recent findings suggest that this bias may also reflect cognitive decline. Specifically, greater positivity bias has been linked to reduced cognitive performance and structural and functional differences in regions such as the anterior hippocampus and amygdala, pointing to its possible role as a marker of early neurodegeneration (Wolpe et al., 2025). Within this context, socially meaningful cues may ‘bypass’ behavioral disengagement and trigger residual autonomic capacity, as indeed found in patients with more advanced dementia and high apathy levels (Treusch et al., 2015). The observed HRV response to positive (grandchildren) images— despite reduced gaze engagement—may thus reflect preserved affective responsiveness selectively activated by socially salient inputs.

These findings carry practical implications. The presence of HRV reactivity despite behavioral disengagement suggests a masked capacity for emotional response. On this account, behavioral apathy can reflect a raised threshold for behavioral engagement that can be selectively overcome with the appropriate stimuli. Clinically, this highlights the potential of leveraging socially salient or personalized stimuli, such as images of family members, to re-engage motivational systems in apathy or cognitive decline. Supporting this notion, interventions incorporating affectively rich content, including reminiscence-based VR or socially tailored environments, have shown great promise in improving engagement in older adults with dementia (Appel et al., 2022). Moreover, physiological reactivity to such stimuli—particularly HRV—may serve as a scalable biomarker for identifying individuals most likely to benefit from these approaches (Manser et al., 2021). More broadly, our findings support the use of naturalistic VR gaze paradigms as a tool for identifying apathy through spontaneous behavior. This approach offers an ecologically valid and scalable alternative to conventional assessments — particularly when combined with autonomic indices such as HRV.

### Strengths and Limitations

The strengths of our study include the use of a VR paradigm specifically designed to assess apathy through spontaneous gaze behavior. Additional strengths include the use of unsupervised clustering without a priori assumptions, and integration of clinical and physiological data. Our results were robust to several control analyses: To reduce the risk of overfitting—an inherent concern in unsupervised models—we used a small and interpretable feature set; validated cluster stability and confirmed robustness via reanalysis after excluding edge-case trials.

However, several limitations should be acknowledged. First, the relatively modest sample size limits statistical power and generalizability. Second, while our aim was to discover behavioral structure relevant to apathy, the lack of a ‘ground truth’ for apathy limits formal cluster validation. Instead, we conducted *post hoc* comparisons with accepted clinical measures of apathy (LARS) for internal validation. However, we acknowledge that this approach does not constitute external validation, and that future studies will be needed to assess the stability and clinical relevance of these clusters longitudinally, as well as the establishment of clinical norms. Third, in terms of physiological analysis, HRV was computed from brief 10-second epochs, which are acceptable for the time-domain index we used but may lack stability compared to longer recordings. Nevertheless, the fact that no significant differences were found in heart rate suggests that the observed differences in HRV reflect true variability rather than random fluctuations in heart rate. Fourth, our study did not fully disentangle the contributions of cognitive impairment and apathy to the observed behavioral patterns. While we controlled for global cognitive function using the MoCA, further research is needed to understand how these two factors independently and interactively influence autonomic reactivity. Future studies with larger longitudinal samples and more granular cognitive assessments may help clarify this relationship.

## Conclusion

In this study, unsupervised clustering of gaze behavior during a naturalistic VR task designed to objectively assess apathy revealed distinct subgroups of older adults differing in clinical features and physiological reactivity. A cluster with reduced behavioral engagement was linked to higher apathy, and lower cognitive scores. Physiologically, the reduced engagement cluster showed reduced autonomic reactivity overall, but a specific increase in reactivity to positive stimuli. These findings highlight the potential of passive, objective behavioral measures for the diagnosis of motivational functioning, and may inform future research on personalized assessment and treatment strategies in psychiatric populations in general, and older adults in particular.

## Data Availability

All data produced in the present study are available upon reasonable request to the authors

## Acknowledgements

NW was supported by an Israel Science Foundation Personal Research Grant (1603/22).

## Conflict of interest

All authors (DH, RRS, SZ, OK, MP, and NW) declare no conflict of interest in relation to the subject of this study.

## Funding

This study was funded by the Alzheimer’s Disease Discovery Foundation (ADDF)-grant number 201906 and by Tel Aviv University Brecher Banner and Ofer Mordechai grants

---

**Supplementary Video 1.** Demonstration of the virtual reality task used in this study, showing the immersive environment and emotional stimuli presented during the session. The video is available at: https://static.psyact.org/harlev_vr/Harlev_etal_apathy_vr.mp4

## Supplementary Information

### Virtual reality task and stimuli

Participants were immersed in a three-dimensional virtual environment developed in Unity3D and presented via an HTC VIVE head-mounted display. The environment included both stationary billboards and moving bus advertisements, which served as platforms for presenting emotionally valenced images (positive, negative and neutral) embedded within a realistic urban park setting. This combination of static and dynamic visual elements and the range of stimuli were selected to elicit variability in emotional and attentional engagement across valence conditions. Neutral stimuli included day-to-day objects such as chairs, while negative stimuli depicted aversive scenes such as bleeding organs or motor vehicle accidents. Aversive and neutral stimuli were selected from validated affective image databases, namely the International Affective Picture System (IAPS) and the Nencki Affective Picture System (NAPS) (Lang et al., 2005; Marchewka et al., 2014). Unknown to participants, their informants provided pictures of their grandchildren for the emotionally positive condition, as we assumed that these are socially meaningful and emotionally salient stimuli (Glocker et al., 2009). A demonstration of the VR stimuli is shown in Supplementary Video 1.

To simulate naturalistic viewing conditions and assess gaze engagement across varying contexts, participants were seated and instructed to freely observe the environment without any explicit task demands Each participant viewed 36 stimuli in total: 12 positive, 12 negative, and 12 neutral images. Each stimulus was displayed for a maximum of 10 seconds—a duration selected to capture gaze and physiological responses while minimizing participant fatigue and aligning with relevant signal frequencies.

### Gaze and physiological measures

Eye-tracking data were recorded using the integrated Pupil Labs system (Pupil Player, Germany) at a sampling rate of 200 Hz. Two gaze metrics were extracted: Time to First Fixation (TTFF) and Total Fixation Duration (TFD), calculated relative to stimulus onset. Physiological signals, including galvanic skin response (GSR), electrocardiogram (ECG) and respiration (rate and depth), were acquired using the EEGO-Sport system (ANT Neuro, The Netherlands) at a sampling rate of 512 Hz. GSR data were pre-processed using the Ledalab toolbox (Benedek & Kaernbach, 2010), and heart rate variability (HRV) indices were computed from ECG signals using the Pan-Tompkins algorithm (Pan & Tompkins, 1985).

To index heart rate variability (HRV), we used the root mean square of successive differences between beats (RMSSD), a time-domain metric that captures rapid beat-to-beat fluctuations primarily driven by parasympathetic activity. RMSSD was selected based on its established validity in ultra-short-term recordings of 10 seconds—consistent with the design and temporal resolution of the current VR task (Shaffer & Ginsberg, 2017). Physiological reactivity was defined as the change in signal amplitude between baseline and post-stimulus periods, using the following formula:

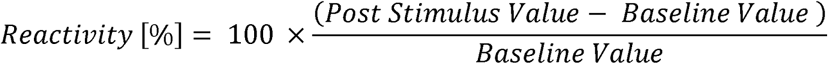

Where baseline values were defined as the 10 seconds preceding stimulus onset, and post-stimulus values were calculated from the 10 seconds following stimulus presentation. Stimuli presentations were separated by a fixed interstimulus interval of 30 seconds in order to allow sufficient time for physiological signals to return to baseline. The five physiological measures (GSR, HR, HRV, respiration rate, and respiration depth) were first pre-processed to detect within-subject outliers, which likely reflect artifacts, using a robust criterion of three scaled median absolute deviations from the median, as done previously (Leys et al., 2013). Following outlier removal, all six physiological variables were scaled using Min-Max normalization to a [0–1] range.

### Unsupervised Clustering Analysis

K-means clustering was applied to three gaze features: mean TTFF, mean TFD, and the Fixation Variability Index (FVI), which was defined as the average of the within-subject standard deviations of TTFF and TFD across all valid trials. TTFF and TFD reflect the speed and duration of attentional allocation, respectively, and were chosen for their relevance to engagement. The mean values of TTFF and TFD were computed across all 36 stimuli. The FVI was, FVI reflects intra-individual variability in gaze behavior, and was chosen because of prior evidence linking increased variability in attentional allocation to apathy-related traits, and an interest in capturing richer within-participant information to enhance the granularity of the clustering space (Yu et al., 2022).

The optimal number of K-means clusters was determined using the Silhouette Score and Davies-Bouldin Index, which are two complementary internal validation metrics for unsupervised clustering (Arbelaitz et al., 2013). The Silhouette Score evaluates clustering quality by measuring the cohesion within clusters and the separation between them, while the Davies-Bouldin Index quantifies cluster compactness and distinctiveness (Ros et al., 2023). To assess the stability of the clustering, we computed the Adjusted Mutual Information (AMI) between the baseline clustering labels and the labels obtained after ‘perturbing’ the dataset with a Gaussian noise (Vinh et al., 2009). This procedure was repeated 1000 times, and AMI values were averaged to quantify consistency in cluster labelling. To ensure that clustering was not affected by the 10-second trial duration limit, in a sensitivity analysis, we excluded trials where [TTFF + TFD] ≥ 9.9 seconds and re-applied the clustering. Consistency in cluster assignments were computed using the Adjusted Rand Index (ARI), which quantifies the agreement between the original and re-clustered labels, corrected for chance.

## Notes

### Competing Interest Statement

The authors have declared no competing interest.

### Author Declarations

The study did not use openly available human data. All data were collected under institutional IRB approval (Sheba Medical Center, IRB #4436-17), and contain identifiable behavioral and physiological information. Due to ethical and privacy restrictions, the data are not publicly available. However, access to de-identified, encrypted datasets may be considered upon reasonable request and subject to approval by the corresponding author and institutional ethics board.

## References

Alfalahi, H., Dias, S. B., Khandoker, A. H., Chaudhuri, K. R., & Hadjileontiadis, L. J. (2023). A scoping review of neurodegenerative manifestations in explainable digital phenotyping. Npj Parkinson’s Disease, 9(1), 1–22. 10.1038/s41531-023-00494-0

Appel, L., Appel, E., Kisonas, E., Lewis, S., & Sheng, L. Q. (2022). Virtual reality for veteran relaxation: Can VR therapy help veterans living with dementia who exhibit responsive behaviors? Frontiers in Virtual Reality, 2, 724020.

Appelhans, B. M., & Luecken, L. J. (2006). Heart Rate Variability as an Index of Regulated Emotional Responding. Review of General Psychology, 10(3), 229–240. 10.1037/1089-2680.10.3.229

Arbelaitz, O., Gurrutxaga, I., Muguerza, J., Pérez, J. M., & Perona, I. (2013). An extensive comparative study of cluster validity indices. Pattern Recognition, 46(1), 243–256.

Beauchaine, T. P., & Thayer, J. F. (2015). Heart rate variability as a transdiagnostic biomarker of psychopathology. International Journal of Psychophysiology, 98(2), 338–350.

Benedek, M., & Kaernbach, C. (2010). A continuous measure of phasic electrodermal activity. Journal of Neuroscience Methods, 190(1), 80–91.

Benjamini, Y., & Yekutieli, D. (2001). The Control of the False Discovery Rate in Multiple Testing under Dependency. The Annals of Statistics, 29(4), 1165–1188.

Bijl, D., Van Marwijk, H. Wj., Adér, H. J., Beekman, A. T. F., & De Haan, M. (2006). Test-Characteristics of the GDS-15 in Screening for Major Depression in Elderly Patients in General Practice. Clinical Gerontologist, 29(1), 1–9. 10.1300/J018v29n01_01

Bock, M. A., Bahorik, A., Brenowitz, W. D., & Yaffe, K. (2020). Apathy and risk of probable incident dementia among community-dwelling older adults. Neurology, 95(24). 10.1212/WNL.0000000000010951

Bufano, P., Laurino, M., Said, S., Tognetti, A., & Menicucci, D. (2023). Digital phenotyping for monitoring mental disorders: Systematic review. Journal of Medical Internet Research, 25, e46778.

Chirico, A., & Gaggioli, A. (2019). When Virtual Feels Real: Comparing Emotional Responses and Presence in Virtual and Natural Environments. *Cyberpsychology*, Behavior, and Social Networking, 22(3), 220–226. 10.1089/cyber.2018.0393

Civitello, D., Finn, D., Flood, M., Salievski, E., Schwarz, M., & Storck, Z. (2014). How do physiological responses such as respiratory frequency, heart rate, and galvanic skin response (GSR) change under emotional stress? https://minds.wisconsin.edu/handle/1793/80044

Egger, M., Ley, M., & Hanke, S. (2019). Emotion recognition from physiological signal analysis: A review. Electronic Notes in Theoretical Computer Science, 343, 35–55.

Glocker, M. L., Langleben, D. D., Ruparel, K., Loughead, J. W., Gur, R. C., & Sachser, N. (2009). Baby Schema in Infant Faces Induces Cuteness Perception and Motivation for Caretaking in Adults. Ethology, 115(3), 257–263. 10.1111/j.1439-0310.2008.01603.x

Harlev, D., Singer, S., Goldshalger, M., Wolpe, N., & Bergmann, E. (2025). Acoustic speech features are associated with late-life depression and apathy symptoms: Preliminary findings. *Alzheimer’s & Dementia (Amsterdam*, Netherlands), 17(1), e70055. 10.1002/dad2.70055

Harlev, D., Vituri, A., Shahar, M., Cam-CAN, & Wolpe, N. (2025). From dysphoria to anhedonia: Age-related shift in the link between cognitive and affective symptoms (p. 2025.03.26.25324666). medRxiv. 10.1101/2025.03.26.25324666

Husain, M., & Roiser, J. P. (2018). Neuroscience of apathy and anhedonia: A transdiagnostic approach. Nature Reviews Neuroscience, 19(8), 470–484.

Kennedy, B. L., & Mather, M. (2024). Negative images, regardless of task relevance, distract younger more than older adults. Psychology and Aging. https://psycnet.apa.org/record/2024-99125-001

Kim, N. (2024). Capturing Initial Gaze Attraction in Branded Spaces Through VR Eye-Tracking Technology. International Journal of Human–Computer Interaction, 1–14. 10.1080/10447318.2024.2351717

Lanctôt, K. L., Ismail, Z., Bawa, K. K., Cummings, J. L., Husain, M., Mortby, M. E., & Robert, P. (2023). Distinguishing apathy from depression: A review differentiating the behavioral, neuroanatomic, and treatmentLrelated aspects of apathy from depression in neurocognitive disorders. International Journal of Geriatric Psychiatry, 38(2), e5882. 10.1002/gps.5882

Lang, P. J., Bradley, M. M., & Cuthbert, B. N. (2005). International affective picture system (IAPS): Affective ratings of pictures and instruction manual. NIMH, Center for the Study of Emotion & Attention Gainesville, FL.

Leys, C., Ley, C., Klein, O., Bernard, P., & Licata, L. (2013). Detecting outliers: Do not use standard deviation around the mean, use absolute deviation around the median. Journal of Experimental Social Psychology, 49(4), 764–766.

Manser, P., Thalmann, M., Adcock, M., Knols, R. H., & de Bruin, E. D. (2021). Can Reactivity of Heart Rate Variability Be a Potential Biomarker and Monitoring Tool to Promote Healthy Aging? A Systematic Review With Meta-Analyses. Frontiers in Physiology, 12. 10.3389/fphys.2021.686129

Marchewka, A., Żurawski, Ł., Jednoróg, K., & Grabowska, A. (2014). The Nencki Affective Picture System (NAPS): Introduction to a novel, standardized, wide-range, high-quality, realistic picture database. Behavior Research Methods, 46(2), 596–610. 10.3758/s13428-013-0379-1

Marshall, G. A., Donovan, N. J., Lorius, N., Gidicsin, C. M., Maye, J., Pepin, L. C., Becker, J. A., Amariglio, R. E., Rentz, D. M., Sperling, R. A., & Johnson, K. A. (2013). Apathy Is Associated With Increased Amyloid Burden in Mild Cognitive Impairment. The Journal of Neuropsychiatry and Clinical Neurosciences, 25(4), 302–307. 10.1176/appi.neuropsych.12060156

Mather, M., & Carstensen, L. L. (2003). Aging and Attentional Biases for Emotional Faces. Psychological Science, 14(5), 409–415. 10.1111/1467-9280.01455

Miller, D. S., Robert, P., Ereshefsky, L., Adler, L., Bateman, D., Cummings, J., DeKosky, S. T., Fischer, C. E., Husain, M., Ismail, Z., Jaeger, J., Lerner, A. J., Li, A., Lyketsos, C. G., Manera, V., Mintzer, J., Moebius, H. J., Mortby, M., Meulien, D., … Lanctôt, K. L. (2021). Diagnostic criteria for apathy in neurocognitive disorders. Alzheimer’s & Dementia, 17(12), 1892–1904. 10.1002/alz.12358

Mohammad, D., Ellis, C., Rau, A., Rosenberg, P. B., Mintzer, J., Ruthirakuhan, M., Herrmann, N., & Lanctôt, K. L. (2018). Psychometric Properties of Apathy Scales in Dementia: A Systematic Review. Journal of Alzheimer’s Disease, 66(3), 1065–1082. 10.3233/JAD-180485

Nasreddine, Z. S., Phillips, N. A., Bédirian, V., Charbonneau, S., Whitehead, V., Collin, I., Cummings, J. L., & Chertkow, H. (2005). The Montreal Cognitive Assessment, MoCA: a brief screening tool for mild cognitive impairment. Journal of the American Geriatrics Society, 53(4), 695–699.

Pan, J., & Tompkins, W. J. (1985). A real-time QRS detection algorithm. IEEE Transactions on Biomedical Engineering, 3, 230–236.

Parsons, T. D. (2015). Virtual reality for enhanced ecological validity and experimental control in the clinical, affective and social neurosciences. Frontiers in Human Neuroscience, 9, 660.

Pedregosa, F., Varoquaux, G., Gramfort, A., Michel, V., Thirion, B., Grisel, O., Blondel, M., Prettenhofer, P., Weiss, R., & Dubourg, V. (2011). Scikit-learn: Machine learning in Python. The Journal of Machine Learning Research, 12, 2825–2830.

Radakovic, R., Harley, C., Abrahams, S., & Starr, J. M. (2015). A systematic review of the validity and reliability of apathy scales in neurodegenerative conditions. International Psychogeriatrics, 27(6), 903–923. 10.1017/S1041610214002221

RavonaLSpringer, R., Koren, O., Galor, N., Lapid, M., Bahat, Y., Fluss, R., Wilf, M., Zorani, S., Rosenblum, U., Beeri, M. S., & Plotnik, M. (2025). A novel method for objective quantification of apathy based on gaze and physiological reactivity to stimuli presented in a virtual reality environment. *Alzheimer’s & Dementia: Diagnosis*, Assessment & Disease Monitoring, 17(1), e70020. 10.1002/dad2.70020

Ros, F., Riad, R., & Guillaume, S. (2023). PDBI: A partitioning Davies-Bouldin index for clustering evaluation. Neurocomputing, 528, 178–199.

Shaffer, F., & Ginsberg, J. P. (2017). An overview of heart rate variability metrics and norms. Frontiers in Public Health, 5, 258.

Sockeel, P., Dujardin, K., Devos, D., Deneve, C., Destée, A., & Defebvre, L. (2006). The Lille apathy rating scale (LARS), a new instrument for detecting and quantifying apathy: Validation in Parkinson’s disease. *Journal of Neurology*, Neurosurgery & Psychiatry, 77(5), 579–584.

Steffens, D. C., Fahed, M., Manning, K. J., & Wang, L. (2022). The neurobiology of apathy in depression and neurocognitive impairment in older adults: A review of epidemiological, clinical, neuropsychological and biological research. Translational Psychiatry, 12(1), Article 1. 10.1038/s41398-022-02292-3

Thant, T., & Yager, J. (2019). Updating Apathy: Using Research Domain Criteria to Inform Clinical Assessment and Diagnosis of Disorders of Motivation. The Journal of Nervous and Mental Disease, 207(9), 707. 10.1097/NMD.0000000000000860

Treusch, Y., Page, J., van der Luijt, C., Beciri, M., Benitez, R., Stammler, M., & Marcar, V. L. (2015). Emotional reaction in nursing home residents with dementia-associated apathy: A pilot study. Geriatric Mental Health Care, 3(1), 1–6. 10.1016/j.gmhc.2015.04.001

Vinh, N. X., Epps, J., & Bailey, J. (2009). Information theoretic measures for clusterings comparison: Is a correction for chance necessary? Proceedings of the 26th Annual International Conference on Machine Learning, 1073–1080. 10.1145/1553374.1553511

Wolpe, N., Harlev, D., Bergmann, E., & Henson, R. (2025). Age-related differences in emotion recognition are linked to cognition and brain structure and function. https://osf.io/tx8dj_v1/

Yu, Y., Xu, H., Xu, Y., Lu, F., & Li, M. (2022). Increased intra-individual variability as a marker of executive dysfunction in generalized anxiety disorder. Frontiers in Psychiatry, 13, 532778.

